# Emergency Department Presenting Concerns Among Admissions With Hypercapnia: A Retrospective NLP Study of MIMIC-IV

**DOI:** 10.64898/2026.07.03.26357242

**Authors:** Reyan Merdad, Monique Ramirez, Matthias Christenson, Warren W. Pettine, Brian W. Locke

**Author notes:** **Corresponding Author** Brian W Locke, MD MSCI, Shock Trauma ICU, Intermountain Medical Center. 5121 Cottonwood Street, Murray Utah.

## Abstract

**Background:** Hypercapnia may indicate a primary ventilatory syndrome, a complication of another illness, or an epiphenomenon of severe disease. The presenting context of hypercapnia is poorly quantified, limiting clinical interpretation and synthesis of epidemiologic studies.

**Methods:** We performed a retrospective cross-sectional study of Medical Information Mart for Intensive Care IV (MIMIC-IV) hospital admissions linked to an emergency department (ED) presentation from 2011 through 2019. Admissions were included if the triage chief complaint was not missing and at least one prespecified criterion for hypercapnia was met: an International Classification of Diseases (ICD) code for hypercapnic respiratory failure or obesity hypoventilation syndrome, arterial blood gas (ABG) PCO ≥45 mmHg, venous blood gas (VBG) PCO ≥50 mmHg, or indeterminate-source blood gas PCO ≥50 mmHg. Triage chief-complaint text was classified by natural language processing (NLP) into 17 National Hospital Ambulatory Medical Care Survey reason-for-visit (RFV) categories using a multi-label framework. Primary analyses estimated admission-level RFV category prevalences; secondary analyses compared distributions by overlapping ascertainment indicator, age, and acidemia.

**Results:** The total cohort included 11,941 admissions: 1,542 (12.9%) met both blood-gas and ICD-code criteria, 9,958 (83.4%) met blood-gas criteria only, and 441 (3.7%) met ICD-code criteria only. Median age at admission was 68 years (IQR 56–78), and 6,423 admissions (53.8%) were for male patients. Respiratory RFV categories were most prevalent (30.2%), followed by administrative reasons (17.5%), digestive symptoms (14.0%), injuries and adverse effects (14.0%), and nervous-system symptoms (13.8%); categories were not mutually exclusive. Respiratory categories were more common in ICD-positive admissions (50.2%) than in VBG-defined (36.3%) or ABG-defined admissions (27.3%). Injuries and adverse effects were most prevalent among admissions for patients aged 18–39 years (34.4%), whereas respiratory categories increased from 13.7% among admissions for patients aged 18–39 years to 36.5% among admissions for patients aged ≥80 years. NLP-derived classifications showed mean set-F1 of 0.84 against adjudicated clinician labels in the full annotated benchmark sample.

**Conclusions:** Among ED-linked admissions with hypercapnia by diagnosis code, blood gas, or both, respiratory complaints were the most common chief-complaint category but represented fewer than one-third of admissions. Presentation context should be incorporated when defining, comparing, and interpreting hypercapnia cohorts, particularly those ascertained by blood-gas criteria.

**Trial registration:** *Not applicable* (observational, no intervention)

## Background

Hypercapnia is a physiologic finding that may indicate the presence of a primary ventilatory syndrome, an important complication of another illness, or an epiphenomenon of severe disease. (1) Yet the ED presentation context of admissions with EHR evidence of hypercapnia has not been quantified, limiting clinicians’ ability to contextualize the finding and complicating interpretation of the epidemiologic literature on any-cause hypercapnia.(2)

Most prior studies of hypercapnic respiratory failure have focused on prevalence, underlying causes (2), ventilatory support (3), readmissions, and mortality (4,5), often in intensive care (6) or other selected cohorts, rather than on how hospital presentation is initially framed. Existing work also relies on non-equivalent physiologic, procedural, and administrative case definitions, which may identify materially different cohorts. (7) As a result, the clinical circumstances surrounding hospital presentation remain poorly described.

Emergency department chief complaints provide a pragmatic record of how illness is initially framed at presentation, before diagnostic testing and final attribution are complete. (8) Their free-text format has traditionally limited systematic study at scale, but natural language processing enables scalable normalization and categorization of these data. (9)

In this retrospective cross-sectional study, we used NLP to classify ED triage chief complaints into standardized National Hospital Ambulatory Medical Care Survey (NHAMCS) reason-for-visit (RFV) categories among MIMIC-IV ED-linked hospital admissions with chief complaint and EHR evidence of hypercapnia by blood gas and/or hypercapnia-related ICD codes. (10,11) We sought to describe ED chief-complaint categories in this EHR-ascertained cohort and to assess how chief-complaint distributions varied.

## Methods

### Study Design and Data Source

We conducted a retrospective cross-sectional study to describe concerns documented in the emergency department triage chief complaint field among MIMIC-IV ED-linked hospital admissions with nonmissing triage chief complaint and EHR evidence of hypercapnia by blood gas and/or hypercapnia-related ICD codes, and to compare concern distributions by overlapping ascertainment indicator, age, and acidemia. We used de-identified data from the Medical Information Mart for Intensive Care IV (MIMIC-IV) database. Emergency department encounters were obtained from MIMIC-IV-ED and linked to corresponding inpatient hospitalizations using unique hospital admission identifiers. MIMIC-IV contains records from Beth Israel Deaconess Medical Center in Boston, Massachusetts, from 2011 through 2019. (10) The MIMIC-IV database was approved by the Institutional Review Boards of the Massachusetts Institute of Technology and Beth Israel Deaconess Medical Center. Because the present analysis used de-identified data, it was not considered human subjects research and did not require additional institutional review board oversight.

### Cohort

We included hospital admissions linked to an emergency department presentation if the triage chief complaint was not missing and at least one prespecified ascertainment criterion for hypercapnia was met: (1) an International Classification of Diseases, Tenth Revision, Clinical Modification (ICD-10-CM) diagnosis code (11) for acute respiratory failure with hypercapnia (J96.02), chronic respiratory failure with hypercapnia (J96.12), acute and chronic respiratory failure with hypercapnia (J96.22), respiratory failure unspecified with hypercapnia (J96.92), or obesity hypoventilation syndrome / morbid obesity with alveolar hypoventilation (E66.2), or ICD-9 code 278.03 (12); (2) any arterial blood gas with PCO ≥45 mmHg; (3) any venous blood gas with PCO ≥50 mmHg (13); or (4) any other blood-gas source with PCO ≥50 mmHg when source classification was indeterminate. When more than one emergency department stay was linked to the same hospital admission, the earliest eligible stay was selected for analysis. We also derived a timing indicator for whether the earliest qualifying hypercapnic gas occurred within 24 hours of emergency department arrival.

The unit of analysis was the hospital admission linked to an ED encounter. Patients could contribute more than one admission; therefore, estimates describe admission-level chief-complaint distributions. Because triage chief complaint is recorded at ED arrival, whereas qualifying blood gases could occur later during hospitalization, the broad cohort was interpreted as hypercapnia identified at any point during the hospital encounter rather than uniformly at ED presentation. We therefore defined timing-safeguard cohorts with first qualifying gas within 24 hours and within 6 hours of ED arrival, plus ICD-positive and ICD-positive with gas within 24 hours cohorts. Separately, we audited availability of pH from the same specimen/panel as the earliest qualifying PCO. Because paired qualifying-gas pH was near-complete, Figure 4 uses paired qualifying-gas pH when available, with source-priority pH retained only as a denominator/context audit. Acidemia figures report analyzed denominators and excluded counts/reasons where pH timing or pH availability is incomplete.

### Variables Extracted

We extracted admission-level variables reported in Table 1, including age, sex, race, triage acuity, ascertainment criteria, selected comorbidities, hospital length of stay, ICU length of stay, mechanical ventilation, and in-hospital death. We also extracted all available arterial, venous, and indeterminate-source blood-gas results from central laboratory and point-of-care data.

**Table 1:** Baseline characteristics of the analytic cohort. Values are shown as median [IQR] or n (%). Percentages use the column denominator unless otherwise specified. The analytic cohort included hospital admissions linked to an emergency department presentation with a nonmissing triage chief complaint and at least one prespecified ascertainment criterion for hypercapnia. Gas-only, ICD-only, and both ICD + gas are mutually exclusive ascertainment strata; ABG-positive, VBG-positive, and ICD-positive are overlapping ascertainment indicators and were not mutually exclusive. ABG, arterial blood gas; CHF, congestive heart failure; COPD, chronic obstructive pulmonary disease; ED, emergency department; ICD, International Classification of Diseases; ICU, intensive care unit; LOS, length of stay; OHS, obesity hypoventilation syndrome; OSA, obstructive sleep apnea; RF, respiratory failure; VBG, venous blood gas.

|  |  |  |  |  |  |
| --- | --- | --- | --- | --- | --- |
| **Demographics** |  |  |  |  |  |
| Age, years | median [IQR] | 68.0 [56.0-78.0] | 67.0 [55.0-78.0] | 66.0 [55.0-76.0] | 69.0 [60.0-79.0] |
| Male sex | n (%) | 6423 (53.8%) | 5565 (55.9%) | 167 (37.9%) | 691 (44.8%) |
| Race |  |  |  |  |  |
| White | n (%) | 7525 (63.0%) | 6375 (64.0%) | 242 (54.9%) | 908 (58.9%) |
| Black or African American | n (%) | 1938 (16.2%) | 1465 (14.7%) | 123 (27.9%) | 350 (22.7%) |
| Other / Unknown | n (%) | 2478 (20.8%) | 2118 (21.3%) | 76 (17.2%) | 284 (18.4%) |
| **ED acuity and ascertainment** |  |  |  |  |  |
| ED triage acuity: 1 | n (%) | 3119 (26.1%) | 2485 (25.0%) | 102 (23.1%) | 532 (34.5%) |
| ED triage acuity: 2 | n (%) | 5482 (45.9%) | 4525 (45.4%) | 202 (45.8%) | 755 (49.0%) |
| ED triage acuity: 3 | n (%) | 1985 (16.6%) | 1658 (16.6%) | 127 (28.8%) | 200 (13.0%) |
| ED triage acuity: 4 | n (%) | 10 (0.1%) | 9 (0.1%) | 1 (0.2%) | 0 (0.0%) |
| ED triage acuity: 5 | n (%) | 1 (0.0%) | 1 (0.0%) | 0 (0.0%) | 0 (0.0%) |
| ED triage acuity missing/unknown | n (%) | 1344 (11.3%) | 1280 (12.9%) | 9 (2.0%) | 55 (3.6%) |
| **Comorbidities** |  |  |  |  |  |
| Chronic obstructive pulmonary disease | n (%) | 2179 (18.2%) | 1351 (13.6%) | 130 (29.5%) | 698 (45.3%) |
| Obstructive sleep apnea/obesity hypoventilation syndrome | n (%) | 1453 (12.2%) | 820 (8.2%) | 163 (37.0%) | 470 (30.5%) |
| Congestive heart failure | n (%) | 3170 (26.5%) | 2192 (22.0%) | 183 (41.5%) | 795 (51.6%) |
| Neuromuscular disease | n (%) | 82 (0.7%) | 53 (0.5%) | 5 (1.1%) | 24 (1.6%) |
| Opioid/substance-related diagnosis | n (%) | 528 (4.4%) | 400 (4.0%) | 22 (5.0%) | 106 (6.9%) |
| Pneumonia | n (%) | 2009 (16.8%) | 1400 (14.1%) | 58 (13.2%) | 551 (35.7%) |
| **Clinical outcomes** |  |  |  |  |  |
| Hospital length of stay, days | median [IQR] | 8.8 [4.8-15.8] | 8.9 [4.9-16.0] | 4.4 [2.8-7.7] | 9.0 [5.0-16.6] |
| ICU length of stay, days | median [IQR] | 3.1 [1.6-6.7] | 3.1 [1.6-6.8] | 1.7 [0.9-3.2] | 3.1 [1.6-6.7] |
| Invasive mechanical ventilation | n (%) | 6725 (56.3%) | 5792 (58.2%) | 84 (19.0%) | 849 (55.1%) |
| Non-invasive ventilation | n (%) | 5155 (43.2%) | 4163 (41.8%) | 80 (18.1%) | 912 (59.1%) |
| Any mechanical ventilation | n (%) | 7626 (63.9%) | 6306 (63.3%) | 133 (30.2%) | 1187 (77.0%) |
| In-hospital death | n (%) | 2110 (17.7%) | 1802 (18.1%) | 26 (5.9%) | 282 (18.3%) |

For subgroup analyses, we used two ascertainment summaries. Ascertainment indicators were overlapping binary indicators for ICD-positive, ABG-positive, and VBG-positive admissions; admissions could meet more than one indicator. Ascertainment strata were mutually exclusive cohort-level groups: gas-only, ICD-only, and both ICD + gas. Age was grouped as 18–39, 40–64, 65–79, and ≥80 years. Among blood-gas-ascertained admissions with paired qualifying-gas pH available, acidemia severity was classified as normal/compensated (pH ≥7.35), mild (7.30–7.34), moderate (7.25–7.29), or severe (<7.25) using pH from the same specimen/panel.(14,15) Acidemia timing was based on the minimum pH across all blood-gas sources within prespecified windows from emergency department arrival: early acidemia (0–6 h pH <7.35), late acidemia (0–6 h pH ≥7.35 and 0–24 h pH <7.35), and no acidemia (0–24 h pH ≥7.35).

### Chief Concern Categorization

The primary outcome was the admission-level prevalence of National Hospital Ambulatory Medical Care Survey (NHAMCS) reason-for-visit (RFV) categories derived from patient-reported concerns documented in the emergency department triage chief complaint field. Concerns were classified into 17 top-level NHAMCS RFV categories spanning 10 symptom-based body-system groups and seven non-symptom categories, including injuries and adverse effects, diseases (patient-stated), abnormal test result, diagnostic/screening/preventive, treatment/medication, administrative, and uncodable/unknown. Primary analyses used a multi-label framework, consistent with NHAMCS conventions that allow up to five RFV assignments per visit; thus, a single admission could contribute to more than one NHAMCS RFV category. As a sensitivity analysis, we repeated summaries using only the first-priority RFV assignment as a mutually exclusive primary-concern classification. The main stratified figures used six grouped RFV categories derived from the canonical RFV labels. Definitions and representative examples for the RFV categories and six grouped analysis categories are provided in Supplementary Appendix 1.

### Natural Language Processing

Triage chief-complaint text was normalized using lowercasing, abbreviation and context expansion, conservative spelling harmonization, and lemmatization. Spelling harmonization used a guarded SymSpell-based strict mode to reduce lexical variation while minimizing distortion of short clinical text. Normalized text was then segmented into discrete concern fragments, with up to five fragments retained per encounter, and embedded using the pretrained clinical sentence-transformer checkpoint NeuML/bioclinical-modernbert-base-embeddings.(16) Fragment embeddings were compared with a curated prototype bank derived from normalized NHAMCS Appendix II descriptors and investigator-curated exemplar phrases; each RFV category was scored by the maximum cosine similarity across its category-specific prototypes. (17,18) Fragments without a deterministic rule override and with a maximum category similarity <0.40 were classified as uncodable. Deterministic rule-based overrides were applied before abstention for common emergency department shorthand and other high-salience clinical patterns. Additional implementation details, including prototype-bank construction, preprocessing rules, and override dictionaries, are provided in the Supplement and code repository.

### Performance Benchmarks vs. Clinicians

To evaluate classification performance, we used a clinician-annotated reference sample of 160 encounters. Two clinician reviewers independently assigned NHAMCS RFV categories to ED triage chief-complaint text while blinded to model output, using a prespecified annotation guide developed for short triage text, multi-complaint entries, and guide-based normalization/segmentation. Disagreements were adjudicated by a third clinician reviewer. Human inter-reviewer agreement was assessed in the full annotated sample, and NLP performance was benchmarked by directly scoring the full adjudicated chief-complaint sample and joining predictions to adjudication labels by annotation row identifier. Agreement was summarized using encounter-level set-based metrics and category-level agreement statistics, including Cohen’s κ and Gwet’s AC1. (19,20) Detailed annotation rules, matching procedures, agreement metrics, and sample-size considerations are provided in Supplementary Appendix 1.

### Statistical Analysis

We summarized admission and patient characteristics using counts and percentages for categorical variables and medians with interquartile ranges for continuous variables. The primary analysis was descriptive and estimated the overall distribution of NHAMCS RFV categories using the multi-label framework described above; because admissions could contribute to more than one RFV category, category percentages were not mutually exclusive and could exceed 100%. These estimates describe admission-level chief-complaint distributions within an EHR-ascertained cohort, not population-level presentation frequencies among all people with hypercapnia. Secondary descriptive analyses compared category distributions by overlapping ascertainment indicator, age group, acidemia severity, and acidemia timing. To assess the robustness of RFV patterns across alternative ascertainment and timing restrictions, we also compared grouped and canonical RFV prevalence across five sensitivity cohorts: the broad EHR-ascertained cohort, admissions with a first qualifying gas within 24 hours, admissions with a first qualifying gas within 6 hours, ICD-positive admissions, and ICD-positive admissions with a qualifying gas within 24 hours. For the main stratified displays, canonical RFV categories were further collapsed into six grouped categories. As sensitivity analyses, we repeated concern summaries using only the first-priority RFV assignment to impose a mutually exclusive primary-concern classification and repeated primary RFV summaries after restricting to the first eligible admission per patient. NLP performance was evaluated against clinician annotation using encounter-level set-based and category-level agreement metrics, including set-F1, Cohen’s κ, and Gwet’s AC1; inter-reviewer agreement was summarized using the same framework. Because the study was descriptive, we did not perform null-hypothesis tests. We estimated 95% confidence intervals for category prevalences using a nonparametric bootstrap clustered by patient to account for repeated admissions. All analyses were performed in Python; data and code availability are described in the Declarations.

## Results

After restricting to hospital admissions linked to an emergency department presentation with a nonmissing triage chief complaint, the analytic cohort comprised 11,941 admissions that met at least one prespecified ascertainment criterion for hypercapnia (Figure 1). Of these, 11,500 (96.3%) met blood-gas criteria; the mutually exclusive ascertainment strata were gas-only in 9,958 admissions (83.4%), ICD-only in 441 (3.7%), and both ICD + gas in 1,542 (12.9%). Across the overlapping ascertainment indicators, 7,454 admissions (62.4%) were ABG-positive, 6,388 (53.5%) were VBG-positive, and 1,983 (16.6%) were ICD-positive. The median age at admission was 68 years (IQR 56–78), and 6,423 admissions (53.8%) were for male patients. Race was recorded as White in 7,525 admissions (63.0%), Black or African American in 1,938 (16.2%), and unknown or not reported in 2,121 (17.8%). Median hospital length of stay was 8.8 days (IQR 4.8–15.8). Additional cohort characteristics are shown in Table 1.

**Figure 1.**
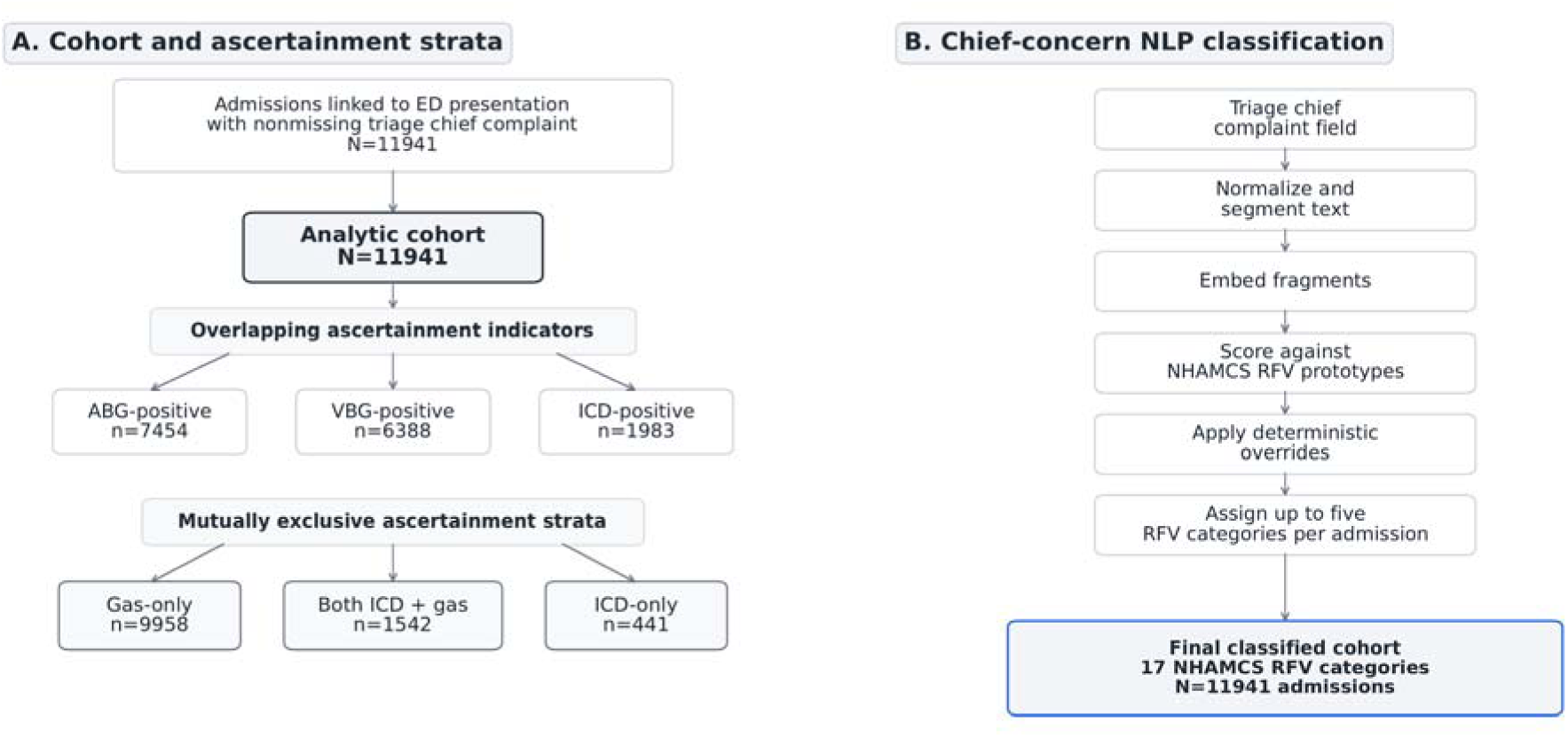
Analytic cohort construction, ascertainment definitions, and chief-concern NLP workflow. Panel A shows construction of the analytic cohort of hospital admissions linked to an emergency department presentation and meeting prespecified administrative or blood-gas criteria for hypercapnia, including the mutually exclusive gas-only, ICD-only, and both ICD + gas ascertainment strata. Panel B shows the natural language processing workflow used to normalize triage chief complaint text, segment presenting-concern fragments, and assign National Hospital Ambulatory Medical Care Survey reason-for-visit categories. Source-specific overlap across ABG-positive, VBG-positive, ICD-positive, and UNKNOWN-source gas ascertainment indicators is shown in Supplementary Figure S6. The primary classification framework was multi-label, allowing up to five RFV assignments per admission. First-priority RFV assignment was used only for sensitivity analyses. ABG, arterial blood gas; ICD, International Classification of Diseases; NLP, natural language processing; RFV, reason for visit; VBG, venous blood gas.

After multi-label NLP classification, respiratory RFV categories were most prevalent (30.2%%; 95% CI, 29.2%–31.2%), followed by administrative reasons (17.5%),; 95% CI, 16.8%–18.3%), digestive symptoms (14.0%; 95% CI, 13.4%–14.7%), injuries and adverse effects (14.0%; 95% CI, 13.4%–14.6%), and nervous-system symptoms (13.8%; 95% CI, 13.2%–14.4%) (Table 2).

**Table 2:** NLP-derived NHAMCS RFV category prevalence. NHAMCS RFV category prevalences are shown after primary multi-label NLP classification. RFV category prevalences are admission-level, not mutually exclusive, and may sum to more than 100% because a single admission could contribute to more than one RFV category. Percentages use the analytic cohort denominator (N = 11941). Confidence intervals are patient-cluster bootstrap 95% confidence intervals. NLP, natural language processing; RFV, reason for visit. Group definitions and examples are provided in Supplementary Appendix 1.

| NHAMCS RFV category | n admissions | % admissions | 95% CI |
| --- | --- | --- | --- |
| Symptom – Respiratory | 3611 | 30.24 | 29.25-31.16 |
| Administrative | 2095 | 17.54 | 16.84-18.27 |
| Symptom – Digestive | 1675 | 14.03 | 13.40-14.72 |
| Injuries & adverse effects | 1672 | 14.0 | 13.36-14.62 |
| Symptom – Nervous | 1651 | 13.83 | 13.19-14.45 |
| Symptom – Circulatory | 1490 | 12.48 | 11.86-13.12 |
| Diseases (patient-stated) | 1327 | 11.11 | 10.53-11.73 |
| Symptom – General | 1225 | 10.26 | 9.72-10.84 |
| Abnormal test result | 495 | 4.15 | 3.75-4.52 |
| Symptom – Musculoskeletal | 433 | 3.63 | 3.31-3.95 |
| Uncodable/Unknown | 311 | 2.6 | 2.32-2.90 |
| Symptom – Skin/Hair/Nails | 253 | 2.12 | 1.85-2.38 |
| Symptom – Genitourinary | 225 | 1.88 | 1.64-2.13 |
| Symptom – Psychological | 132 | 1.11 | 0.92-1.30 |
| Treatment/Medication | 101 | 0.85 | 0.67-1.04 |
| Symptom – Eye/Ear | 49 | 0.41 | 0.30-0.53 |
| Diagnostic/Screening/Preventive | 8 | 0.07 | 0.03-0.12 |

Presenting-concern categories varied by how hypercapnia was identified (Figure 2). Respiratory RFV categories were present in 50.2% (95% CI, 47.8%–52.7%) of ICD-positive admissions, compared with 36.3% (95% CI, 34.9%–37.6%) of VBG-defined admissions and 27.3% (95% CI, 26.2%–28.3%) of ABG-defined admissions. Non-respiratory categories remained common across all ascertainment indicators. Other grouped RFV categories were present in 39.9% of ICD-positive, 44.6% of VBG-defined, and 41.3% of ABG-defined admissions; injuries and adverse effects were present in 8.0%, 11.5%, and 16.3%, respectively.

**Figure 2.**
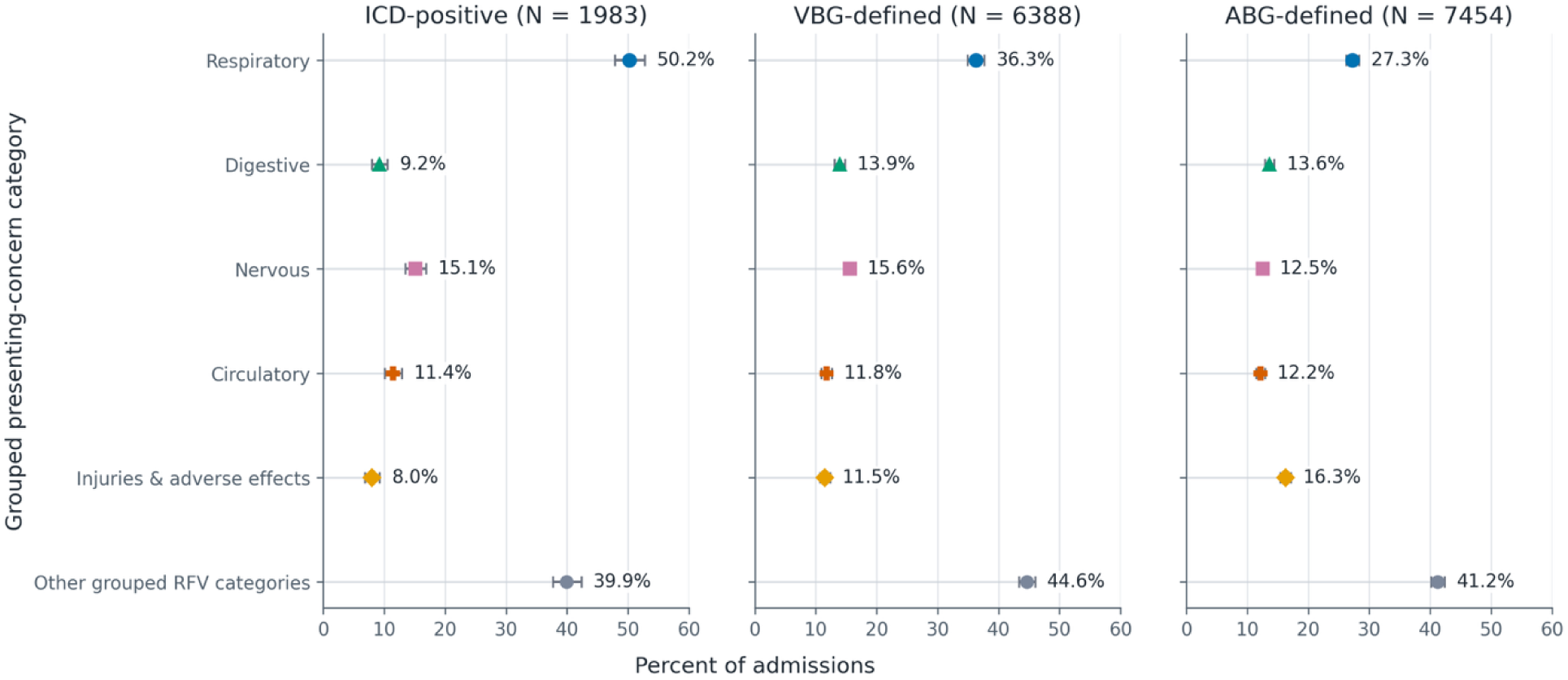
Grouped presenting-concern category prevalence across overlapping ascertainment indicators. Grouped NHAMCS RFV category prevalence is shown across overlapping ascertainment indicators: ICD-positive, VBG-positive, and ABG-positive admissions. Admissions may contribute to more than one panel. Whiskers show patient-cluster bootstrap 95% confidence intervals. Categories were assigned using the primary multi-label RFV framework; therefore, percentages are not mutually exclusive and may sum to more than 100%. Panel denominators are shown in the figure. “Other grouped RFV categories” includes all canonical RFV categories not displayed separately as respiratory, digestive, nervous, circulatory, or injuries and adverse effects. ABG, arterial blood gas; ICD, International Classification of Diseases; RFV, reason for visit; VBG, venous blood gas.

Presenting-concern distributions varied across age and acidemia strata. Injuries and adverse effects were the most prevalent grouped category among admissions for patients aged 18–39 years (34.4%; 95% CI, 31.2%–37.6%), whereas respiratory categories increased from 13.7% (95% CI, 11.1%–16.3%) among admissions for patients aged 18–39 years to 36.5% (95% CI, 34.5%–38.6%) among admissions for patients aged 80 years or older (Figure 3). Across prespecified paired-pH acidemia severity strata, respiratory categories decreased from 33.6% (95% CI, 32.0%–35.1%) in normal/compensated admissions to 23.6% (95% CI, 21.8%–25.6%) in severe acidemia, while injuries and adverse effects increased from 10.3% (95% CI, 9.4%–11.3%) to 18.3% (95% CI, 16.7%–20.0%) (Figure 4). Acidemia timing patterns are shown in Supplementary Figure S1 among admissions with sufficient pH timing data; early acidemia had a higher prevalence of injuries and adverse effects than no acidemia (27.9% vs 11.1%), whereas no-acidemia admissions had a higher prevalence of respiratory categories (35.0% vs 27.4%). The Supplementary source data workbook reports denominator and excluded-count details for pH-timing missingness.

**Figure 3.**
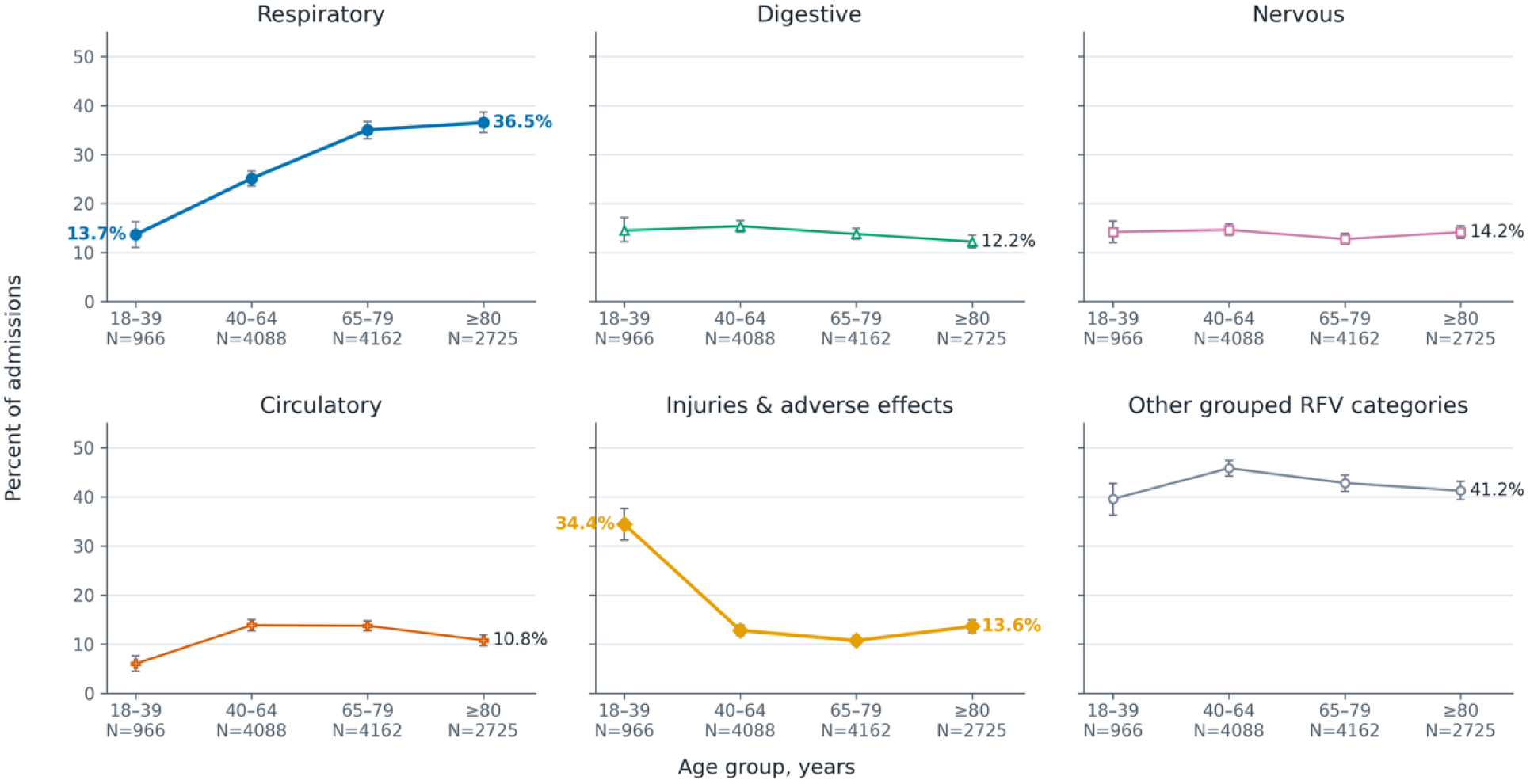
Grouped presenting-concern category prevalence across age groups. Grouped NHAMCS RFV category prevalence is shown across prespecified age strata: 18–39, 40–64, 65–79, and ≥80 years. Whiskers show patient-cluster bootstrap 95% confidence intervals. Categories were assigned using the primary multi-label RFV framework; therefore, percentages are not mutually exclusive and may sum to more than 100%. Panel denominators are shown in the figure. RFV, reason for visit.

**Figure 4.**
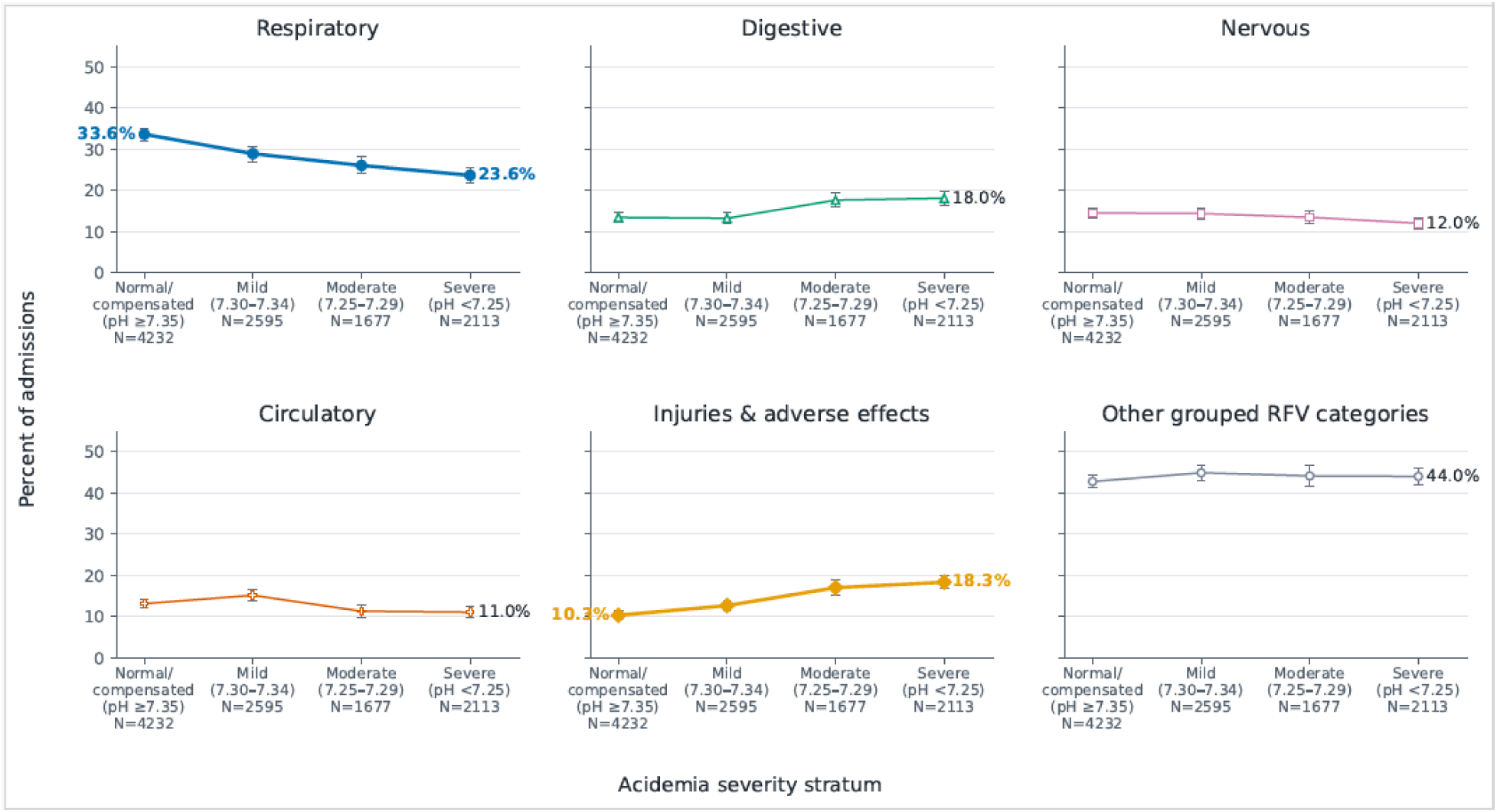
Grouped presenting-concern category prevalence across acidemia severity strata. Grouped NHAMCS RFV category prevalence is shown across prespecified acidemia severity strata among blood-gas-ascertained admissions with pH from the same specimen/panel as the earliest qualifying PCO available. Whiskers show patient-cluster bootstrap 95% confidence intervals. Acidemia severity was classified as normal/compensated (pH ≥7.35), mild (pH 7.30–7.34), moderate (pH 7.25–7.29), or severe (pH <7.25). Categories were assigned using the primary multi-label RFV framework; therefore, percentages are not mutually exclusive and may sum to more than 100%. Panel denominators and paired-pH missingness/provenance counts are shown in the Supplementary source data workbook. RFV, reason for visit.

Early-gas timing restrictions yielded chief-complaint distributions similar to the broad cohort, while ICD-positive admissions showed a more respiratory-concentrated presentation profile. The first qualifying gas occurred within 24 hours in 6,218 admissions (52.1% of the broad cohort) and within 6 hours in 2,654 admissions (22.2%). Respiratory RFV categories were present in 32.8% (95% CI, 31.5%–34.1%) of 24-hour early-gas admissions and 33.8% (95% CI, 31.9%– 35.7%) of 6-hour early-gas admissions, compared with 30.2% (95% CI, 29.2%–31.2%) in the broad cohort; other grouped RFV categories remained the largest group in both early-gas cohorts (39.5% and 38.1%). By contrast, ICD-positive admissions were more respiratory-concentrated (50.2%; 95% CI, 47.8%–52.7%), and admissions with both an ICD phenotype and a qualifying gas within 24 hours had the highest respiratory prevalence (58.5%.; 95% CI, 55.3%–61.5%).

In the clinician-annotated reference sample (n = 160), the two independent clinician reviewers had exact set agreement in 80.6% of encounters and partial agreement in 13.1%; mean set-F1 was 0.88, macro-averaged Cohen’s κ was 0.73, and macro-averaged Gwet’s AC1 was 0.97. In the direct annotation NLP benchmark (n = 160), NLP-derived classifications had exact agreement with the adjudicated reference in 76.3% of encounters and partial agreement in 12.5%; mean set-F1 was 0.84, macro-averaged Cohen’s κ was 0.63, and macro-averaged Gwet’s AC1 was 0.97.

## Discussion

In this retrospective study of ED-linked hospital admissions with evidence of hypercapnia or hypercapnia-related diagnosis codes, respiratory reason-for-visit categories were the most prevalent chief-complaint category overall but were present in fewer than one-third of admissions. The chief-complaint profile differed by overlapping ascertainment indicator: ICD-positive admissions were more concentrated in respiratory categories, whereas blood-gas-ascertained admissions showed a broader distribution of nonrespiratory chief complaints. Similar patterns in early-gas timing-safeguard cohorts suggest that this broad presenting-concern distribution was not driven only by hypercapnia identified later in the hospital encounter. Taken together, the findings suggest that hypercapnia in hospitalized patients spans a wider clinical spectrum than classic ventilatory failure presentations alone.

Prior studies establish that hypercapnia and hypercapnic respiratory failure are heterogeneous and associated with substantial morbidity. Population-based studies have described prevalence, causes, mortality, and rehospitalization, while hospital-based cohorts have focused on ventilatory support, health care use, readmission, and mortality in cohorts identified by blood-gas values, diagnosis codes, or respiratory failure admissions. (12,21) These studies provide important epidemiologic context, but they generally characterize admissions after hypercapnia has been recognized rather than describing the presenting concern that brought the patient to emergency care. Our study adds a complementary perspective by using ED triage concern text to characterize the presentation context surrounding admissions in which hypercapnia is documented or coded. NLP methods have been used to structure ED triage text and support prediction tasks, but, to our knowledge, they have not been used to characterize the chief-complaint spectrum of admissions with EHR-documented hypercapnia. (22–24)

Respiratory RFV categories were more common among diagnosis code–positive admissions, while blood gas–identified admissions showed a wider distribution of presenting concerns. This contrast suggests that diagnosis codes and blood-gas criteria capture related but differently weighted populations: coded admissions emphasize clinically recognized hypercapnic respiratory failure, whereas gas-identified admissions include a broader set of acute illnesses with measured hypercapnia. As a result, the clinical presentation of a hypercapnia cohort will depend partly on the case-definition elements used to define it.

Age and acidemia analyses showed that the presenting-concern distribution also varied across clinically important subgroups. Younger patients more often presented with injuries or adverse effects, while older patients more often presented with respiratory concerns. Across acidemia strata, respiratory concerns were more common in normal or compensated hypercapnia, whereas injury-related presentations were more common in severe acidemia, supporting the broader finding that hypercapnia can appear across different acute-care contexts.

Strengths of this study include the large cohort, the use of both blood-gas results and diagnosis codes to identify admissions with evidence of hypercapnia, and scalable NLP classification of triage concerns with clinician benchmarking. Several limitations warrant discussion The cohort included only hospital admissions linked to an ED encounter, with nonmissing triage chief-complaint text and evidence of hypercapnia from blood-gas results or diagnosis codes. It therefore does not represent all patients with hypercapnia, and ED-only encounters were not included. Because these eligibility criteria depend on healthcare use and documentation, conditioning on them can induce selection and collider bias. (25) A second limitation is that study variables came from routinely collected clinical data rather than protocolized assessment in all patients. Blood-gas results depended on which patients clinicians chose to test. Diagnosis codes reflected clinician recognition and billing documentation rather than a uniform reference standard for respiratory failure. Chief complaints were brief triage fields and may have captured transfer, referral, or administrative workflow rather than the patient’s dominant symptom. For blood gas–identified admissions, the qualifying gas could occur after triage; early-gas timing-safeguard analyses reduced this temporal mismatch but did not eliminate it. Finally, the NLP model was benchmarked against clinician-adjudicated labels in a limited sample, so performance for rare RFV categories may be lower or less precisely estimated than global metrics suggest.

These findings have clinical and methodological implications. For clinical interpretation, they suggest that hypercapnia appears across a broader range of admissions than classic respiratory syndromes alone. Methodologically, these findings show that ICD-based ascertainment captures a more respiratory-concentrated subset than blood-gas ascertainment and may miss admissions in which hypercapnia is documented outside classic respiratory presentations. Triage concern text adds context that blood-gas values and diagnosis codes cannot provide; NLP-based RFV classification offers a scalable way to characterize the presentation context surrounding physiologic findings and administrative labels. Future work should test whether combining physiologic criteria, administrative labels, and presenting-concern phenotypes improves cohort definition and case identification.

## Conclusions

In this EHR cohort of ED-linked admissions with documented hypercapnia or hypercapnia-related diagnosis codes, respiratory complaints were the most common chief-complaint category but accounted for only a minority of admissions. The chief-complaint distribution, particularly among blood-gas–ascertained admissions, suggests that documented hypercapnia includes patients with classic respiratory presentations and patients whose initial presenting concerns are not primarily respiratory. Incorporating presentation context may improve how hypercapnia cohorts are defined, compared, and interpreted.

## List of Abbreviations

ABG: arterial blood gas
AC1: Gwet’s AC1 agreement coefficient
BMI: body mass index
CHF: congestive heart failure
CO: carbon dioxide
COPD: chronic obstructive pulmonary disease
ED: emergency department
F1 score: harmonic mean of precision and recall
ICD: International Classification of Diseases
ICU: intensive care unit
IMV: invasive mechanical ventilation
IRB: Institutional Review Board
IQR: interquartile range
LOS: length of stay
MIMIC-IV: Medical Information Mart for Intensive Care IV
NHAMCS: National Hospital Ambulatory Medical Care Survey
NIV: non-invasive ventilation
NLP: natural language processing
OMR: online medical record
OSA: obstructive sleep apnea
OHS: obesity hypoventilation syndrome
PaCO: arterial partial pressure of carbon dioxide
PvCO: venous partial pressure of carbon dioxide
RF: respiratory failure
VBG: venous blood gas
κ: Cohen’s kappa

## Ethics approval and consent to participate

This study was conducted using de-identified data from the Medical Information Mart for Intensive Care IV (MIMIC-IV) database. The MIMIC-IV database has received prior ethical approval from the Institutional Review Boards of the Massachusetts Institute of Technology and Beth Israel Deaconess Medical Center, with a waiver of informed consent due to the use of de-identified data. This study did not involve direct human subjects research and was therefore exempt from additional institutional review board oversight.

## Consent for publication

Not applicable

## Availability of data and materials

The data that support the findings of this study are available from the MIMIC-IV database (https://physionet.org/), which requires credentialed access. The analytical code used in this study is available at: https://github.com/reblocke/Hypercap-CC-NLP

## Declaration of generative AI and AI-assisted technology use

Generative AI tools were used to support code drafting/debugging and manuscript editing. The authors reviewed and verified all code, outputs, citations, and text and take full responsibility for the final manuscript.

## Authors’ contributions

BL, RM, WP, MR, MC contributed to study conception and design. BL & RM performed data extraction and analysis. RM, MC & BL developed the natural language processing pipeline. BL, RM, MR performed clinician annotation and adjudication. BL & RM drafted the manuscript. All authors contributed to interpretation of data, critically revised the manuscript for important intellectual content, and approved the final version.

## Acknowledgements

The authors would like to acknowledge the contributors to the MIMIC-IV database for providing access to a high-quality, publicly available dataset.

## Tables and Figures

**Supplementary Figure S1.**
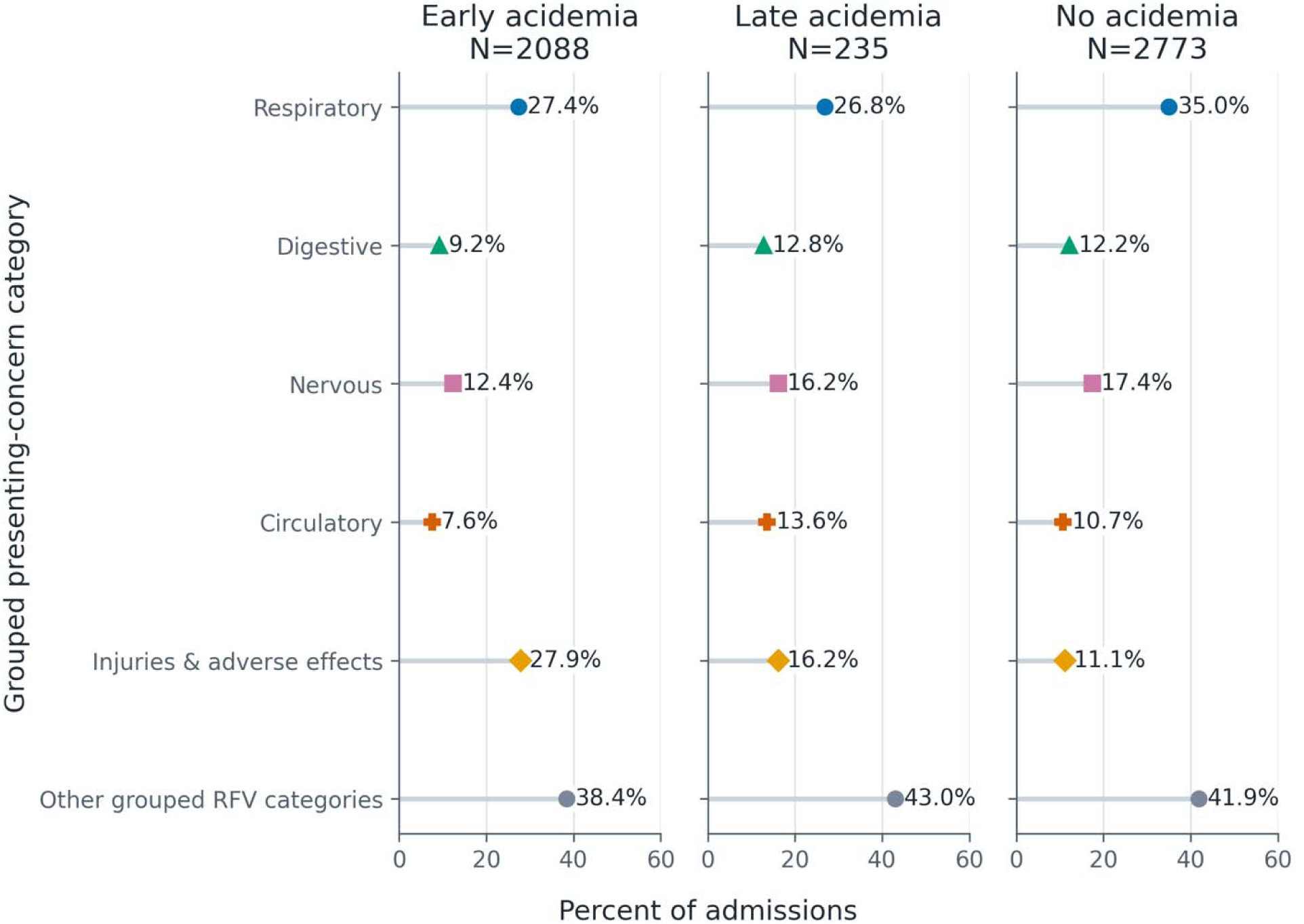
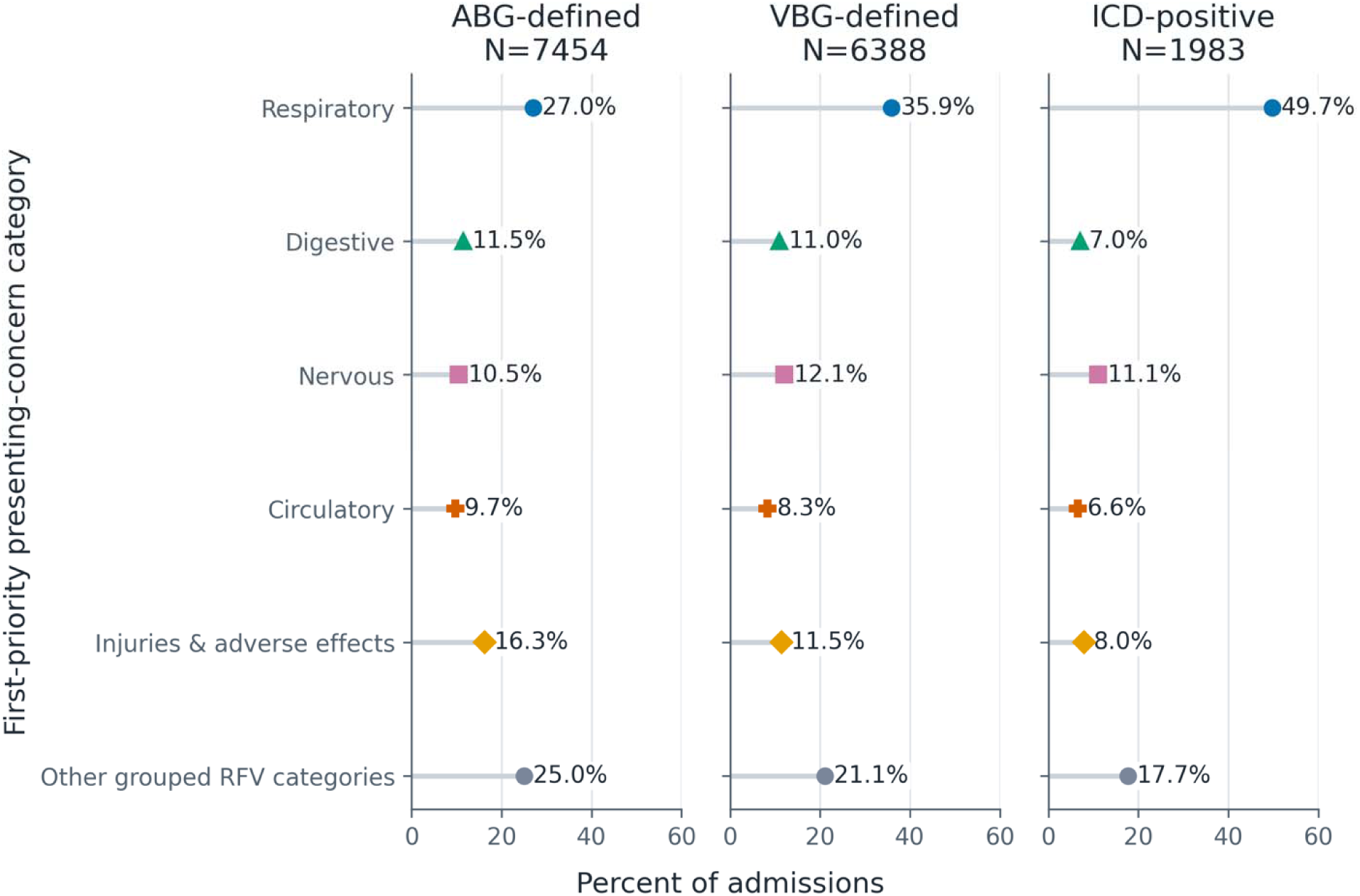
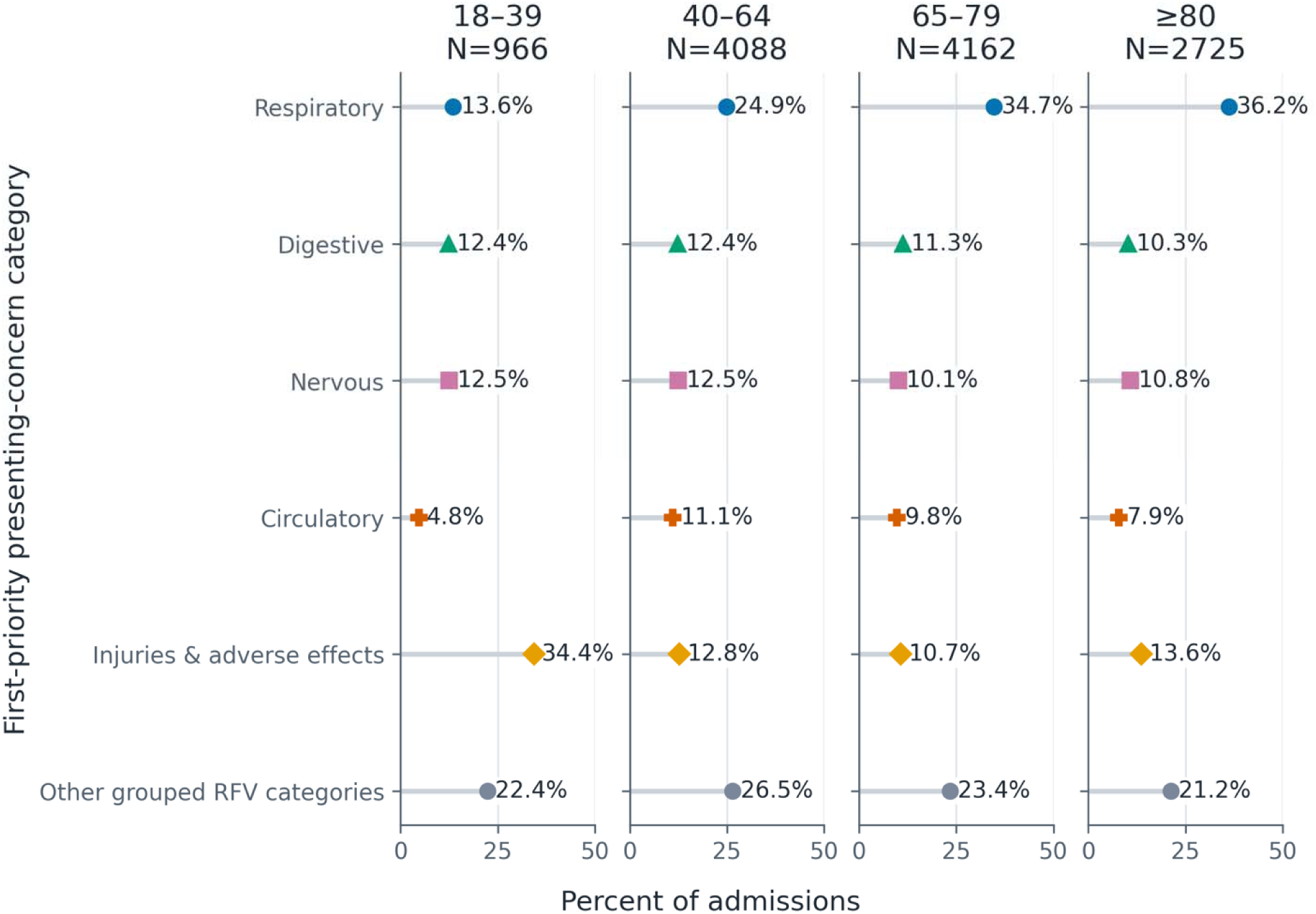
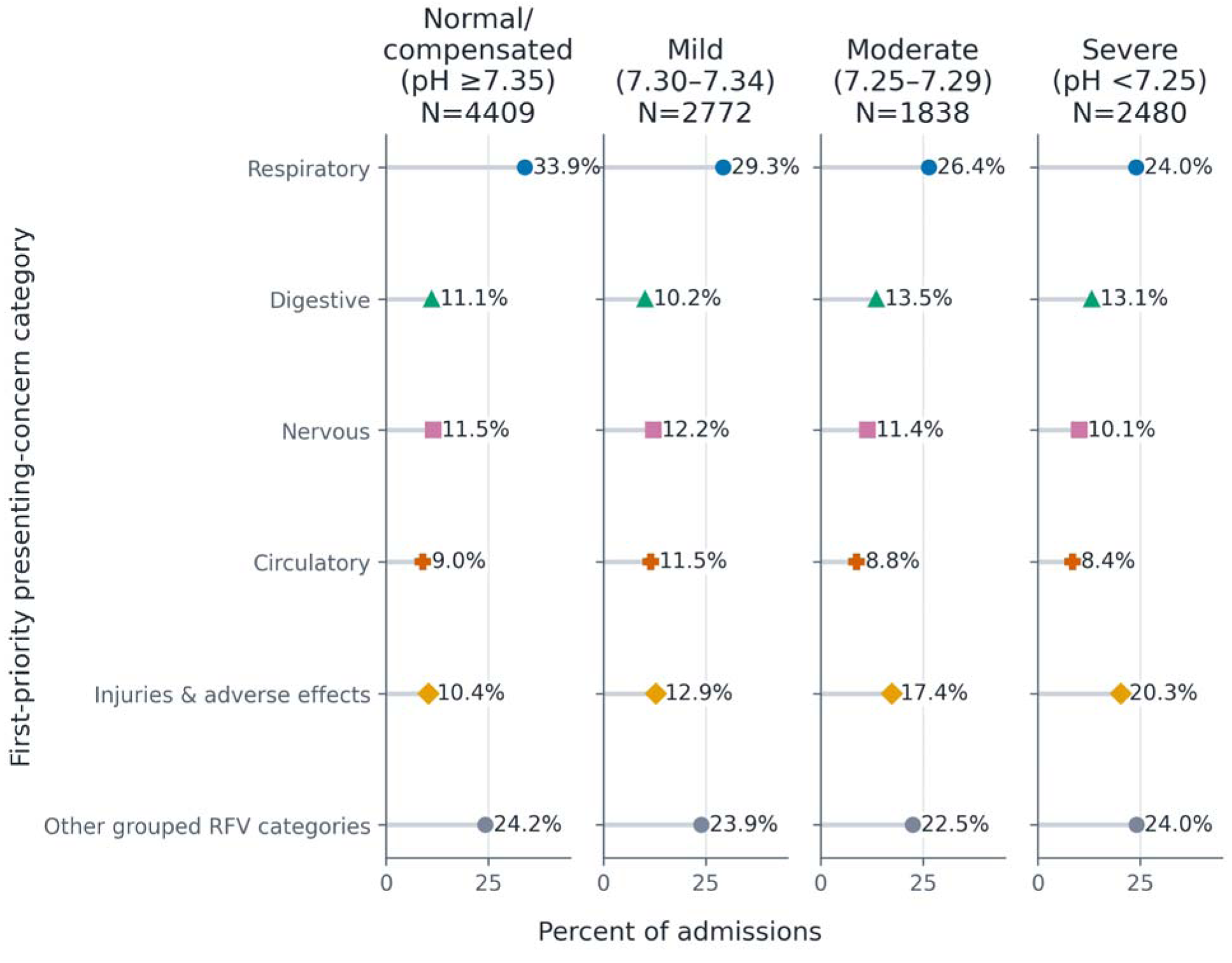
Grouped presenting-concern category prevalence across acidemia timing strata. Grouped NHAMCS RFV category prevalence is shown across early acidemia, late acidemia, and no-acidemia timing strata. Acidemia timing was classified using pH values in prespecified 0–6 h and 0–24 h windows from ED arrival. This analysis is restricted to admissions with qualifying PCO and sufficient pH data in the specified timing windows; the Supplementary source data workbook reports excluded counts and reasons, including no pH within 24 h and acidemia within 24 h without pH in the 0–6 h window. Categories were assigned using the primary multi-label RFV framework; therefore, percentages are not mutually exclusive and may sum to more than 100%. Panel denominators are shown in the figure. RFV, reason for visit.

**Supplementary Figures S2–S5.**
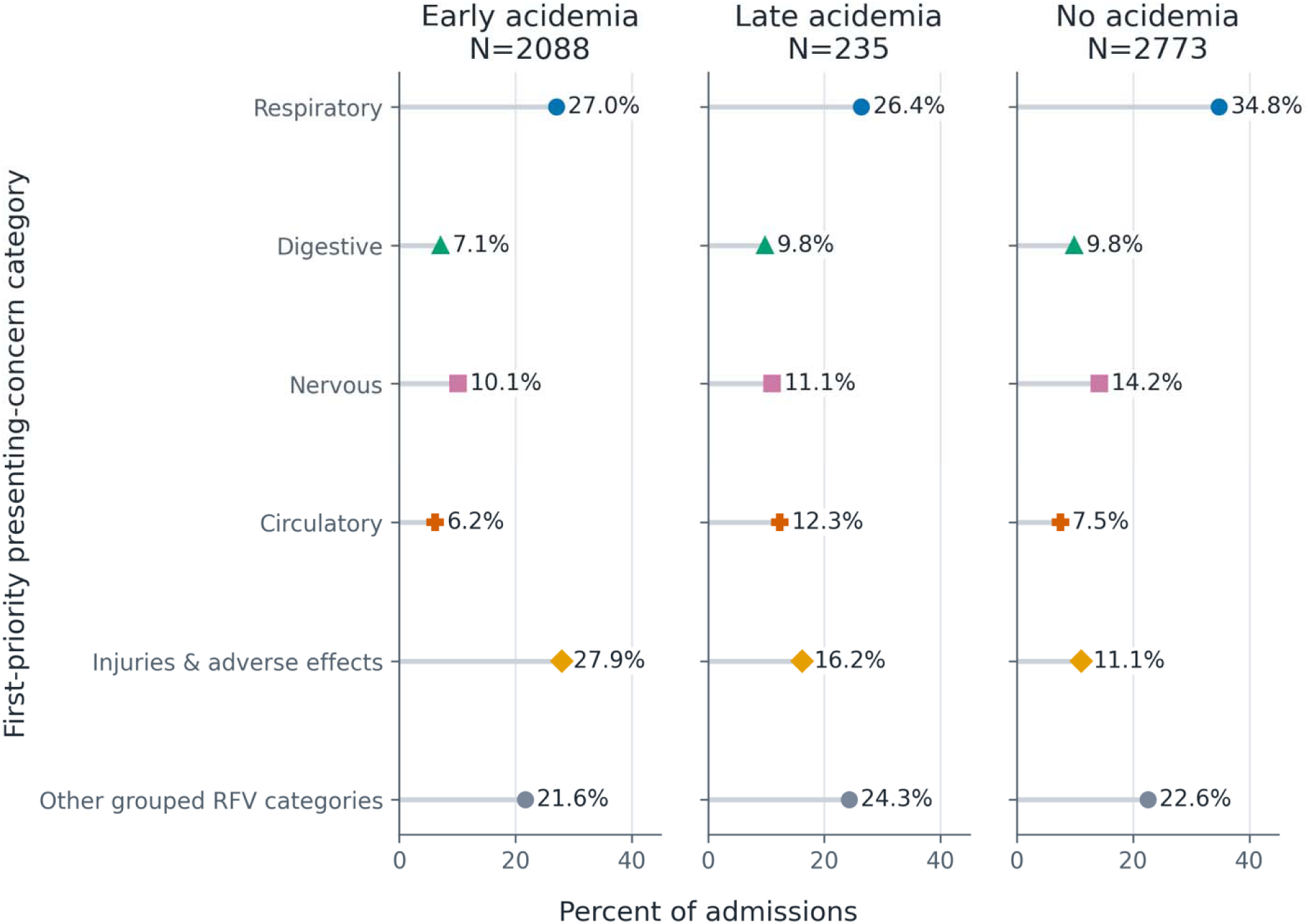
First-priority RFV sensitivity analyses. Supplementary Figures S2–S5 repeat the main grouped RFV displays using only the first-priority RFV assignment as a mutually exclusive sensitivity analysis. Supplementary Figure S2 compares ascertainment indicators, Supplementary Figure S3 compares age groups, Supplementary Figure S4 compares acidemia severity strata, and Supplementary Figure S5 compares acidemia timing strata. RFV, reason for visit.

**Supplementary Figure S6.**
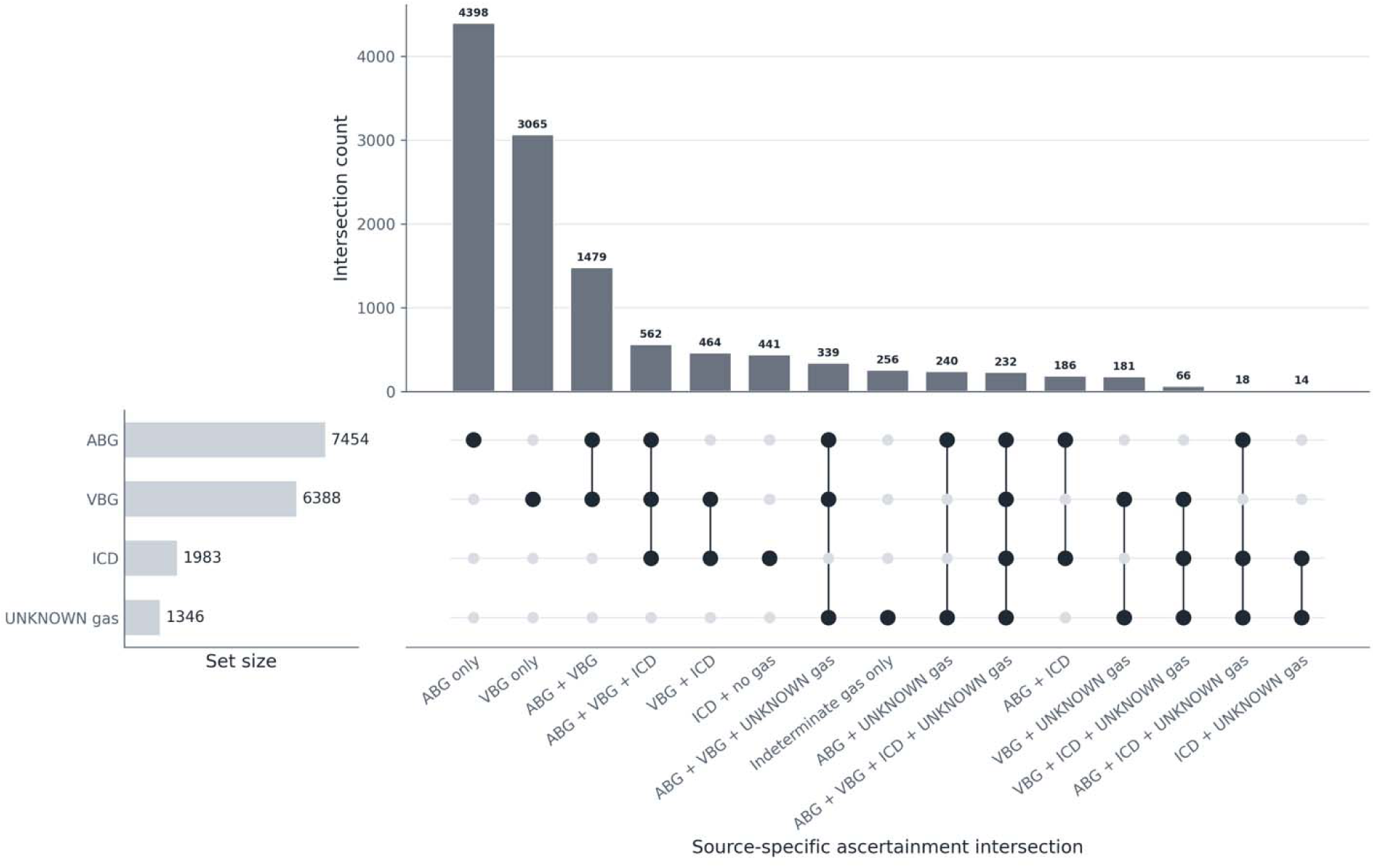
Expanded source-specific ascertainment overlap across ABG, VBG, ICD, and UNKNOWN-source gas. Supplementary UpSet-style source-specific overlap summary showing ordered intersection counts across ABG, VBG, ICD, and UNKNOWN-source gas ascertainment sources. Figure 2 remains limited to ABG/VBG/ICD overlapping ascertainment indicators. ABG, arterial blood gas; ICD, International Classification of Diseases; VBG, venous blood gas.

**Supplementary Figure S7.**
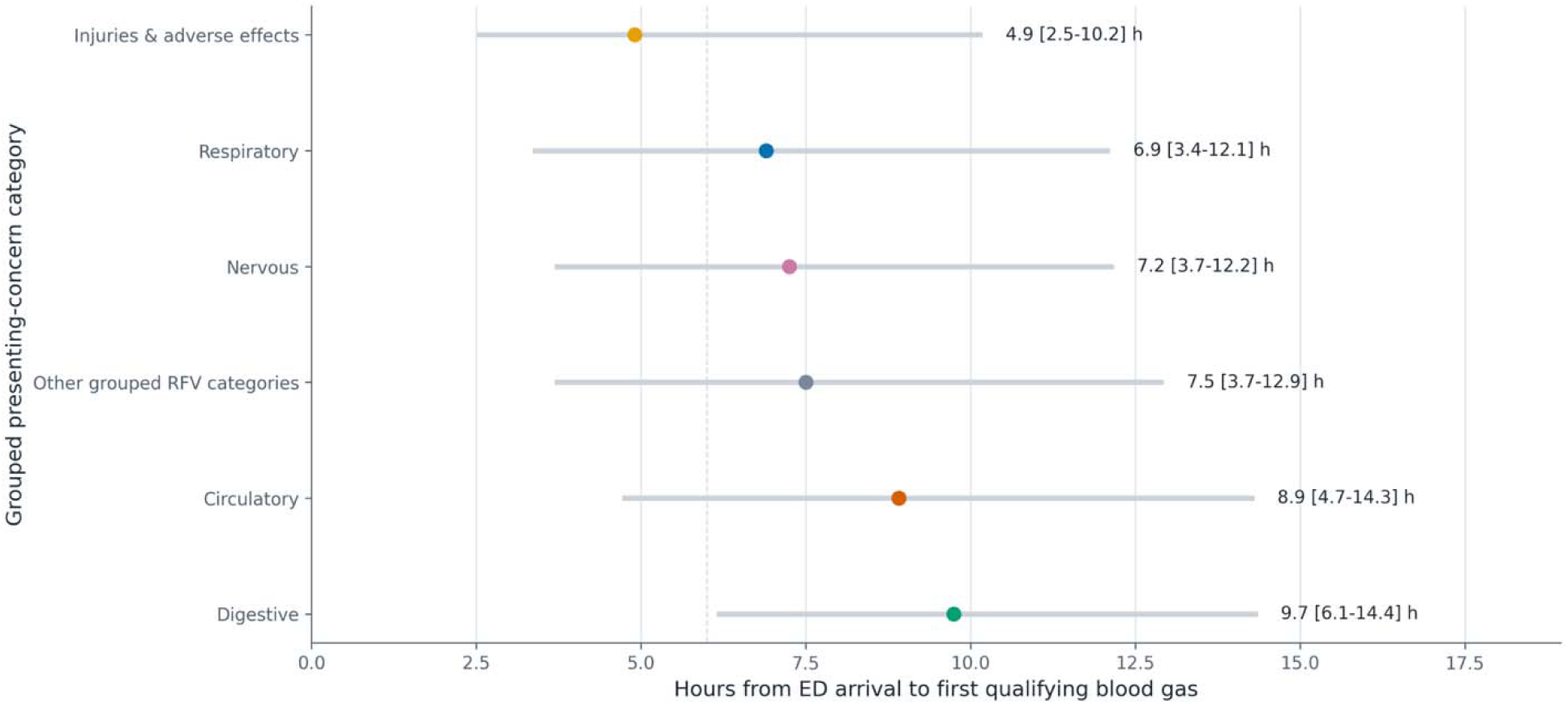
Time to first blood gas by presenting-concern category. Median and interquartile range of time from ED arrival to first qualifying blood gas documentation are shown for grouped presenting-concern categories within the blood-gas subset. This analysis describes timing of blood-gas documentation and should not be interpreted as diagnostic delay. ED, emergency department.

**Supplementary Figure S8.**
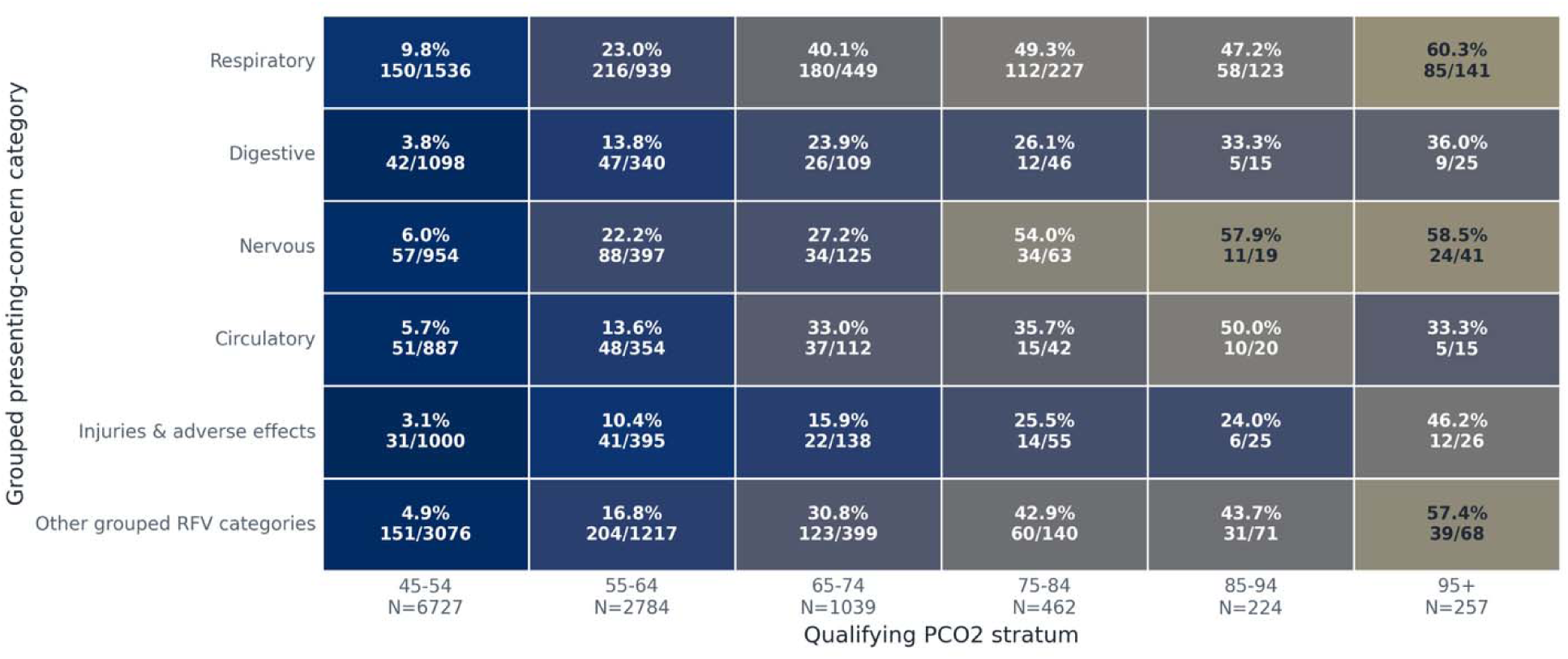
ICD recognition versus pCO2 severity by category. Matrix heatmap showing percent ICD-positive across qualifying pCO2 bins for grouped presenting-concern categories. ICD recognition is descriptive and should not be interpreted as diagnostic accuracy without chart review. ICD, International Classification of Diseases.

**Supplementary Figure S9.**
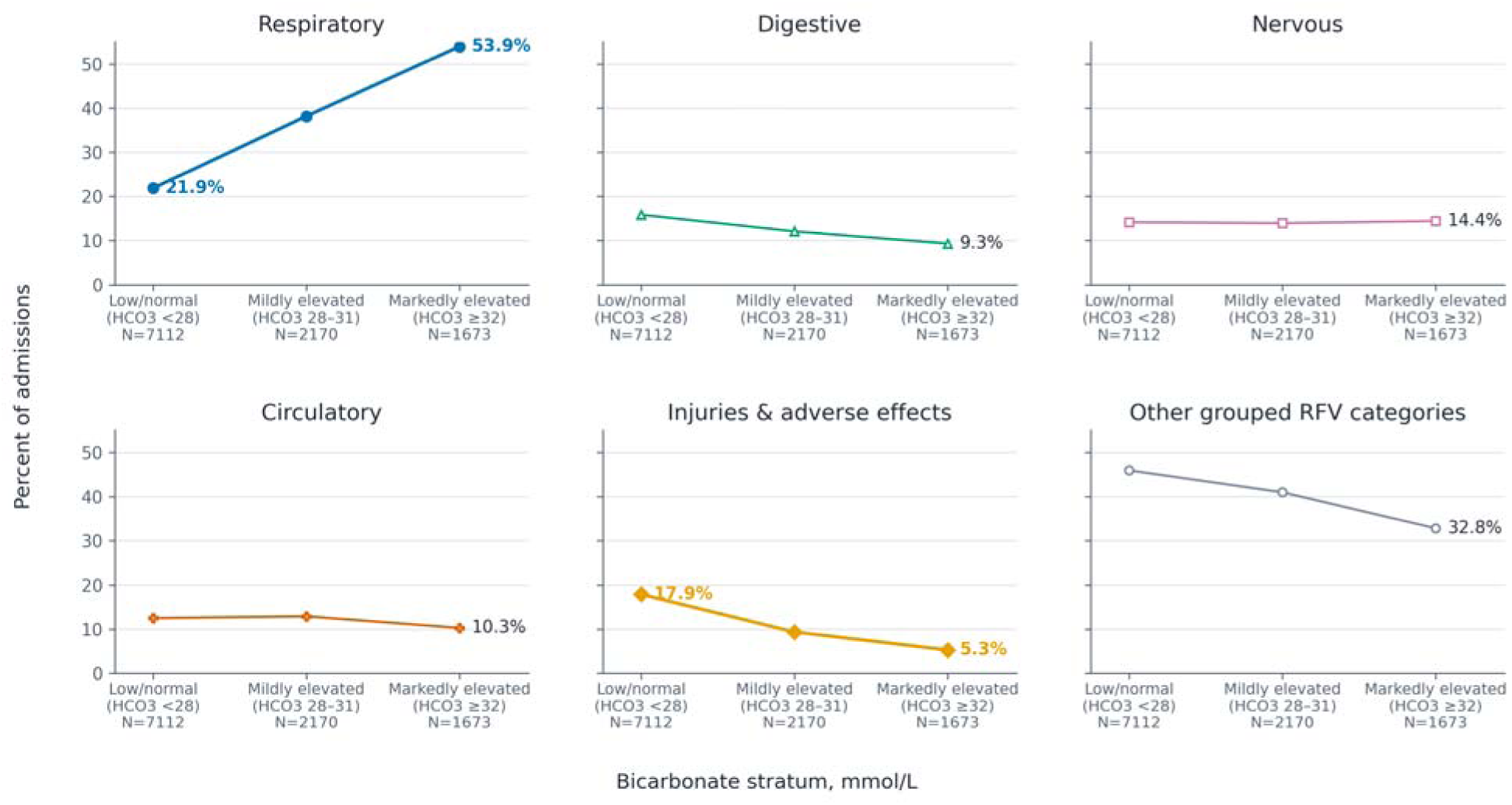
Grouped presenting-concern category prevalence across bicarbonate strata. Grouped admission-level presenting-concern category prevalence is shown across bicarbonate strata among blood-gas-ascertained admissions with available bicarbonate; percentages are multi-label and not mutually exclusive. Bicarbonate strata are presented as descriptive evidence of acid-base compensation and should not be interpreted as definitive chronicity classification.

